# COMMUNITY KNOWLEDGE AND ATTITUDE TOWARDS POLIO IMMUNIZATION IN NORTHERN NIGERIA

**DOI:** 10.64898/2026.09.29.26364257

**Authors:** Muhammad Auwal Gidado, Abar Okoye Ifeoma, Suke Halimat

**Affiliations:** Department of Public Health, College of Health Technology Ningi, Bauchi State, Nigeria; Medical Laboratory Science Council of Nigeria, Abuja, Nigeria; Peace Restoration and Integral Global Development Initiative (PRIDE Initiative), Nigeria

**Keywords:** Poliomyelitis, Oral Polio Vaccine, Vaccine Hesitancy, Northern Nigeria, Community Health, Routine Immunization

## Abstract

**Background:** Poliomyelitis remains a public health concern where population immunity is incomplete and circulating vaccine-derived poliovirus (cVDPV) persists. Although Nigeria interrupted wild poliovirus transmission in 2020, cVDPV cases continue to pose challenges, particularly in parts of Northern Nigeria.

**Methods:** This multicentre descriptive cross-sectional study evaluated community knowledge and attitudes towards polio immunization across 37 Local Government Areas (LGAs) in 20 states and the Federal Capital Territory (FCT). A multistage sampling strategy was utilized to select 1,850 adult respondents (50 per LGA) spanning rural, semi-urban, and urban settlements.

**Results:** Overall, 40.4% (n = 747) of respondents demonstrated good knowledge, while 58.4% (n = 1,080) displayed positive attitudes towards polio immunization. A striking disparity was observed across residence classifications: respondents in urban/semi-urban LGAs exhibited significantly higher levels of good knowledge (51.7%, 569/1,100) compared to those in rural LGAs (23.7%, 178/750; χ^2^ = 144.00, df = 1, p < 0.001). Main sources of information were health workers (33.0%) and radio (21.1%).

**Conclusion:** Knowledge deficits persist in rural Northern Nigerian communities despite favorable attitudes. Interventions must prioritize localized risk communication, traditional and religious leadership involvement, and primary healthcare integration.

## 1. INTRODUCTION

Poliomyelitis is a highly infectious viral disease caused by poliovirus, primarily affecting children under five years of age. The virus invades the central nervous system, leading to acute flaccid paralysis (AFP), irreversible paralysis, respiratory failure, or death in a subset of cases. Because there is no curative medical treatment for paralytic polio, effective prevention relies exclusively on maintaining high population immunity through widespread vaccination coverage and robust epidemiological surveillance.

The Global Polio Eradication Initiative (GPEI) has successfully reduced wild poliovirus (WPV) transmission worldwide by more than 99% since its inception. Nigeria achieved a major public health milestone in August 2020 when it was certified free of wild poliovirus following three consecutive years without detecting WPV type 1. However, the country continues to contend with circulating vaccine-derived polioviruses (cVDPV), notably cVDPV type 2 (cVDPV2) and occasionally cVDPV3. These strains emerge in areas with persistent low routine immunization coverage, where the attenuated oral polio vaccine (OPV) virus circulates long enough to regain neurovirulence.

Socio-demographic disparities in healthcare access and health literacy across Nigeria significantly impact vaccination uptake. According to the 2018 Nigeria Demographic and Health Survey (NDHS), only 31% of children aged 12–23 months received all basic vaccinations, with urban children nearly twice as likely as rural children to be fully immunized (44% vs. 23%). Furthermore, national coverage for the third dose of the polio vaccine (Polio3) stood at 47%, highlighting persistent gaps in completion. Northern Nigeria faces unique structural and social challenges, including geographical inaccessibility, insecurity, population displacement, gendered health decision-making, and lingering historical mistrust stemming from past vaccination boycotts.

Community knowledge, attitudes, and perceptions play a decisive role in vaccine acceptance. Misinformation, low perceived disease risk, fear of adverse events following immunization (AEFI), and institutional distrust remain critical drivers of vaccine hesitancy. Conversely, effective community engagement involving traditional rulers, religious leaders, local health workers, and targeted door-to-door interpersonal communication has proven essential in building trust and overcoming refusal. This study presents a comprehensive multicentre assessment of community knowledge and attitudes towards polio immunization across selected rural, semi-urban, and urban LGAs in Northern Nigeria and the FCT.

## 2. METHODOLOGY

### 2.1 Study Design & Area

A multicentre descriptive cross-sectional study design was utilized across 37 Local Government Areas (LGAs) distributed within 20 States and the Federal Capital Territory (FCT), representing the North East, North West, and North Central geopolitical zones of Nigeria. The target population comprised adult community members (aged 18 years and older), specifically targeting parents and primary caregivers of children eligible for routine polio immunization.

### 2.2 Sample Size Determination & Statistical Formula

To establish an adequate sample size for estimating population proportions with high precision, Cochran’s (1977) sample size formula for cross-sectional study designs was applied:

**Cochran’s Sample Size Formula:**

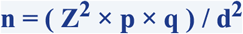

Where: **n** = Target sample size; **Z** = Standard normal deviation at 95% confidence level (1.96); **p** = Estimated proportion of good knowledge/positive attitude (assumed 0.50 for maximum variability); **q** = 1 − p = 0.50; **d** = Margin of error allowable (0.05).

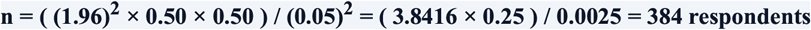

To accommodate multi-stage cluster sampling design effects (DEFF = 1.2) and potential non-response or incomplete questionnaire completion (10%), the operational sample size was allocated across 37 selected LGAs with 50 respondents per LGA, yielding a total study population of N = 1,850 respondents.

### 2.3 Sampling & Data Collection

A multi-stage sampling strategy was employed: Stage 1 selected 20 states and FCT; Stage 2 selected 37 LGAs spanning rural, semi-urban, and urban classifications; Stage 3 selected two wards per LGA; Stage 4 systematically sampled households; and Stage 5 randomly selected one eligible adult caregiver per household. Data were collected using a pre-tested structured interviewer-administered questionnaire.

## 3. RESULTS

**Table 3.1:**
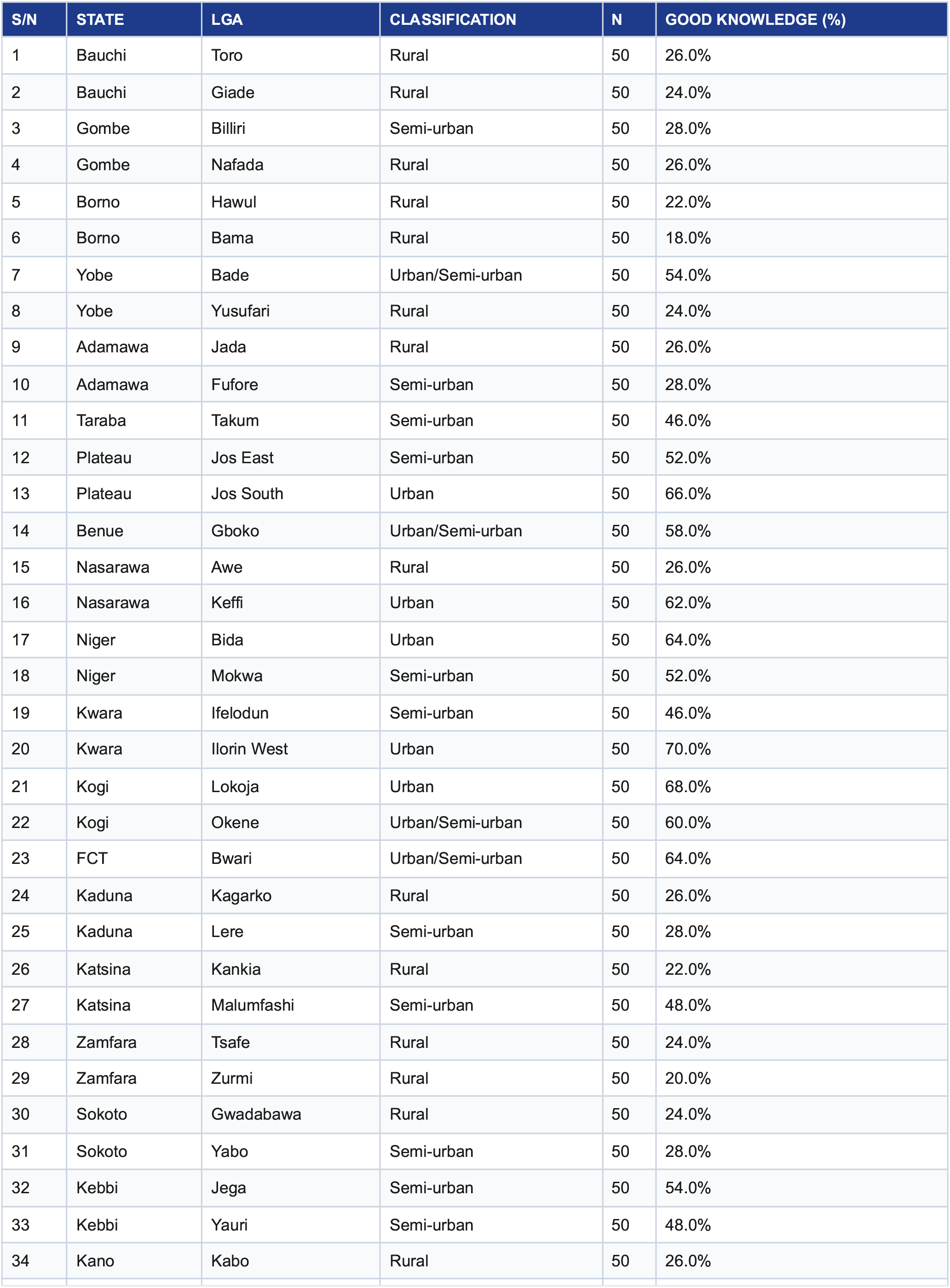
Distribution of Respondents and Knowledge Levels Across Selected LGAs (N = 1,850)

**Table 3.2:**
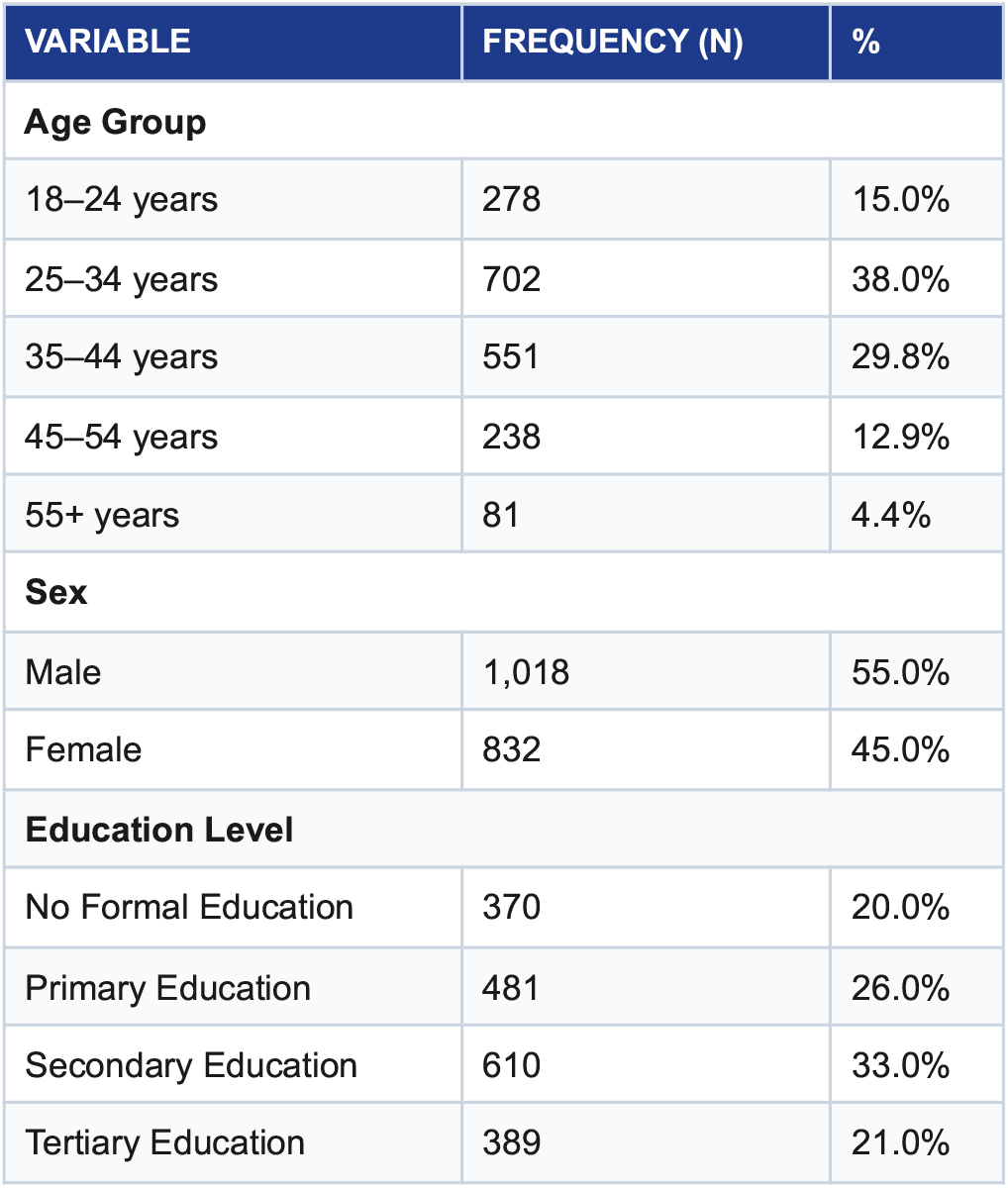
Socio-Demographic Profile (N = 1,850)

| VARIABLE | FREQUENCY (N) | % |
| --- | --- | --- |
| **Age Group** |  |  |
| 18–24 years | 278 | 15.0% |
| 25–34 years | 702 | 38.0% |
| 35–44 years | 551 | 29.8% |
| 45–54 years | 238 | 12.9% |
| 55+ years | 81 | 4.4% |
| **Sex** |  |  |
| Male | 1,018 | 55.0% |
| Female | 832 | 45.0% |
| **Education Level** |  |  |
| No Formal Education | 370 | 20.0% |
| Primary Education | 481 | 26.0% |
| Secondary Education | 610 | 33.0% |
| Tertiary Education | 389 | 21.0% |

Table 3.3: Overall Knowledge & Attitude.

Table 3.3: Overall Knowledge & Attitude
| CATEGORY | FREQUENCY (N) | % |
| --- | --- | --- |
| **Knowledge Status** |  |  |
| Good Knowledge ($\geq 70\%$) | 747 | 40.4% |
| Poor Knowledge ($< 70\%$) | 1,103 | 59.6% |
| **Attitude Category** |  |  |
| Positive Attitude ($\geq 70\%$) | 1,080 | 58.4% |
| Neutral Attitude (50–69%) | 360 | 19.5% |
| Negative Attitude ($< 50\%$) | 410 | 22.1% |

**Table 3.4:** Cross-Tabulation of Knowledge Level by Settlement Setting.

| SETTLEMENT SETTING | GOOD KNOWLEDGE (%) | POOR KNOWLEDGE (%) | TOTAL (N) | CHI-SQUARE (X<sup>2</sup>) | P-VALUE |
| --- | --- | --- | --- | --- | --- |
| Predominantly Rural | 178 (23.7%) | 572 (76.3%) | 750 (100.0%) | 144.00 | < 0.001 |
| Urban / Semi-urban | 569 (51.7%) | 531 (48.3%) | 1,100 (100.0%) |  |  |
| Total Overall | 747 (40.4%) | 1,103 (59.6%) | 1,850 (100.0%) | df = 1 |  |

**Table 3.5:** Information Sources.

| SOURCE | N | % |
| --- | --- | --- |
| Health Workers / Facilities | 610 | 33.0% |
| Radio Broadcasts | 390 | 21.1% |
| Traditional Leaders | 250 | 13.5% |
| Television | 220 | 11.9% |
| Family & Friends | 190 | 10.3% |
| Town Announcers | 120 | 6.5% |
| Social Media / Internet | 70 | 3.7% |

## 4. DISCUSSION & RECOMMENDATIONS

This study demonstrates a significant knowledge deficit regarding polio immunization in rural Northern Nigeria despite a high proportion of positive community attitudes (58.4%). Urban respondents exhibited more than double the rate of good knowledge compared to rural peers (51.7% vs. 23.7%; χ2 = 144.00, p < 0.001). Health workers (33.0%) and radio broadcasts (21.1%) remain the primary trusted drivers of health information.

### Recommendations

1) Intensify localized health education in remote rural LGAs; 2) Systematically engage traditional and religious leaders as key vaccine advocates; 3) Integrate polio campaigns into routine primary healthcare services; 4) Enhance microplanning to reach zero-dose children in mobile and insecure populations.

## Supporting information

Supplementary author list

## Data Availability

All data produced in the present work are contained in the manuscript

## APPENDIX I SURVEY INSTRUMENT & CODING SCHEME

**Selected Questionnaire Items & Variable Dictionary**

| DOMAIN | SURVEY ITEM / STATEMENT | RESPONSE OPTIONS | CODING VALUE |
| --- | --- | --- | --- |
| Knowledge | Is polio a preventable disease? | Yes / No / Don't Know | 1 = Correct (Yes), 0 = Incorrect |
| Knowledge | Can polio cause permanent paralysis in children? | Yes / No / Don't Know | 1 = Correct (Yes), 0 = Incorrect |
| Attitude | Polio vaccination effectively protects children against serious disease. | 5-point Likert Scale | 1 = SD to 5 = SA |
| Attitude | I am concerned that polio vaccines might cause harm. (Reverse) | 5-point Likert Scale | 1 = SA to 5 = SD |
| Access | Have vaccinators visited your household before? | Yes / No / Don't Know | 1 = Yes, 0 = No |
| Outcome | Overall Knowledge Classification | Good vs. Poor | 1 = ≥70% score, 0 = <70% |

**Informed Consent Note:** Verbal and written informed consent was obtained from all respondents prior to questionnaire administration. Participation was strictly voluntary, and no personal identifying information was collected.

## Notes

### Competing Interest Statement

The authors have declared no competing interest.

### Author Declarations

Ethical approval was obtained from the Ministry of Health / Abubakar Tafawa Balewa University Teaching Hospital Bauchi, Health Research Ethics Committee. Informed verbal and written consent were obtained from all participants prior to questionnaire administration. Respondent anonymity and data confidentiality were strictly maintained.

### Summary of Updates

This version has a revised version of author's list

