## Supplementary author list for "COMMUNITY KNOWLEDGE AND ATTITUDE TOWARDS POLIO IMMUNIZATION IN NORTHERN NIGERIA"

**ABSTRACT**

Background: Poliomyelitis remains a major public health concern where population immunity is incomplete and circulating vaccine-derived poliovirus (cVDPV) persists. Although Nigeria interrupted wild poliovirus (WPV) transmission in 2020, cVDPV cases continue to pose challenges, particularly in parts of northern Nigeria. Modern control strategies require a synergy of routine immunization, supplementary campaigns, disease surveillance, and community trust.

Methods: This multicentre descriptive cross-sectional study evaluated community knowledge and attitudes towards polio immunization across 37 Local Government Areas (LGAs) in 20 states and the Federal Capital Territory (FCT). A multistage sampling strategy was utilized to select 1,850 adult respondents (50 per LGA) spanning rural, semi-urban, and urban settlements.

Results: Overall, 40.4% (n = 747) of respondents demonstrated good knowledge, while 58.4% (n = 1,080) displayed positive attitudes towards polio immunization. A striking disparity was observed across residence classifications: respondents in urban/semi-urban LGAs exhibited significantly higher levels of good knowledge (51.7%, 569/1,100) compared to those in rural LGAs (23.7%, 178/750; χ² = 144.00, df = 1, p < 0.001). Main sources of information were Questionnaire responses (53.6%) and community and Traditional leaders(17.2%).

Conclusion & Recommendations: Substantial knowledge deficits persist in rural northern Nigerian communities despite generally favorable attitudes. Interventions must prioritize localized risk communication, involvement of traditional/religious leaders, strengthening routine primary healthcare integration, and targeting zero-dose children in hard-to-reach areas.

**Keywords:** *Poliomyelitis, Oral Polio Vaccine, Vaccine Hesitancy, Northern Nigeria, Community Health, Routine Immunization.*

**1. INTRODUCTION**

Poliomyelitis is a highly infectious viral disease caused by poliovirus, primarily affecting children under five years of age. The virus invades the central nervous system, leading to acute flaccid paralysis (AFP), irreversible paralysis, respiratory failure, or death in a subset of cases. Because there is no curative medical treatment for paralytic polio, effective prevention relies exclusively on maintaining high population immunity through widespread vaccination coverage and robust epidemiological surveillance.

The Global Polio Eradication Initiative (GPEI) has successfully reduced wild poliovirus (WPV) transmission worldwide by more than 99% since its inception. Nigeria achieved a major public health milestone in August 2020 when it was certified free of wild poliovirus following three consecutive years without detecting WPV type 1. However, the country continues to contend with circulating vaccine-derived polioviruses (cVDPV), notably cVDPV type 2 (cVDPV2) and occasionally cVDPV3. These strains emerge in areas with persistent low routine immunization coverage, where the attenuated oral polio vaccine (OPV) virus circulates long enough to regain neurovirulence [2, 4].

Socio-demographic disparities in healthcare access and health literacy across Nigeria significantly impact vaccination uptake. According to the 2018 Nigeria Demographic and Health Survey (NDHS), only 31% of children aged 12–23 months received all basic vaccinations, with urban children nearly twice as likely as rural children to be fully immunized (44% vs. 23%) [6]. Furthermore, national coverage for the third dose of the polio vaccine (Polio3) stood at 47%, highlighting persistent gaps in completion [6]. Northern Nigeria faces unique structural and social challenges, including geographical inaccessibility, insecurity, population displacement, gendered health decision-making, and lingering historical mistrust stemming from past vaccination boycotts [8, 9, 13].

Community knowledge, attitudes, and perceptions play a decisive role in vaccine acceptance. Misinformation, low perceived disease risk, fear of adverse events following immunization (AEFI), and institutional distrust remain critical drivers of vaccine hesitancy [13, 18]. Conversely, effective community engagement involving traditional rulers, religious leaders, local health workers, and targeted door-to-door interpersonal communication has proven essential in building trust and overcoming refusal [7, 10, 17]. This study presents a comprehensive multicentre assessment of community knowledge and attitudes towards polio immunization across selected rural, semi-urban, and urban LGAs in Northern Nigeria and the Federal Capital Territory (FCT).

**2. METHODOLOGY**

**2.1 Study Design**

A multicentre descriptive cross-sectional study design was utilized. This approach is well-suited for assessing knowledge levels, behavioral attitudes, sources of information, and socio-demographic determinants across diverse populations at a single point in time.

**2.2 Study Area and Population**

The study was conducted across 37 Local Government Areas (LGAs) distributed within 20 States and the Federal Capital Territory (FCT), representing the North East, North West, and North Central geopolitical zones of Nigeria. The study population comprised adult community members (aged 18 years and above), specifically targeting parents and primary caregivers of children eligible for routine polio immunization who had resided in the study area for at least six months.

**2.3 Sample Size Determination and Formula**

To establish an adequate sample size for estimating population proportions with high precision, Cochran's (1977) formula for cross-sectional studies was applied:

**n = (Z² × p × q) / d²**

- **n:** Desired target sample size
- **Z:** Standard normal deviation corresponding to a 95% confidence level = 1.96
- **p:** Estimated baseline proportion of good knowledge/positive attitude = 0.50 (assumed 50% for maximum variability)
- **q:** 1 - p = 0.50
- **d:** Margin of error allowable = 0.05 (5%)

**Calculation:** n = ((1.96)² × 0.50 × 0.50) / (0.05)²
n = (3.8416 × 0.25) / 0.0025 = 384.16 ≈ 384 respondents per major zone/stratum.

To accommodate multi-stage cluster sampling design effects (DEFF = 1.2) and potential non-response or incomplete questionnaire completion (10%), the operational sample size was allocated across 37 selected LGAs with 50 respondents per LGA, yielding a total study population of N = 1,850 respondents.

**2.4 Sampling Technique**

A multi-stage sampling approach was employed to select eligible respondents:

- **Stage 1:** Selection of 20 states across the three Northern geopolitical zones and the FCT.
- **Stage 2:** Selection of 37 LGAs based on strategic geographical coverage, routine immunization performance metrics, and settlement type (rural vs. urban/semi-urban).
- **Stage 3:** Random selection of two wards per selected LGA.
- **Stage 4:** Systematic sampling of households within designated ward communities/settlements.
- **Stage 5:** Simple random selection of one eligible adult caregiver per selected household.

**2.5 Data Collection, Management, and Measurement**

Data were collected using a structured, pre-tested interviewer-administered questionnaire (Appendix I) covering socio-demographics, knowledge, attitude, and access factors. Forward and back-translations into local languages (Hausa and Fulani) were performed to ensure conceptual consistency.

**Measurement Rules:**

- **Knowledge:** Evaluated across 10 factual items scored 1 for correct and 0 for incorrect/don't know responses. Total scores were converted to percentages and categorized as Good Knowledge (≥70%) or Poor Knowledge (<70%).
- **Attitude:** Assessed using 12 statements on a 5-point Likert scale (1=Strongly Disagree to 5=Strongly Agree). Scores were summed, with negative statements reverse-scored, and categorized into Positive Attitude (≥70%) vs. Neutral/Negative Attitude (<70%).
- **Analysis:** Cleaned data were analyzed using SPSS version 26. Descriptive statistics (frequencies, percentages, means) and inferential statistics (Pearson's Chi-Square test, logistic regression) were computed at a significance threshold of p < 0.05.

**2.6 Ethical Considerations**

Ethical approval was obtained from relevant institutional review committees. Informed verbal and written consent were obtained from all participants prior to questionnaire administration. Respondent anonymity and data confidentiality were strictly maintained.

**3. RESULTS**

A total of 1,850 respondents completed the structured interview across the 37 LGAs. Table 3.1 outlines the geographical distribution, residence classification, and observed knowledge levels per LGA.

**Table 3.1: Distribution of Respondents and Knowledge Levels by Study Area (N = 1,850)**

| **S/N** | **State** | **LGA** | **Classification** | **n** | **Good Knowledge (%)** |
| --- | --- | --- | --- | --- | --- |
| 1 | Bauchi | Toro | Rural | 50 | 26.0% |
| 2 | Bauchi | Giade | Rural | 50 | 24.0% |
| 3 | Gombe | Billiri | Semi-urban | 50 | 28.0% |
| 4 | Gombe | Nafada | Rural | 50 | 26.0% |
| 5 | Borno | Hawul | Rural | 50 | 22.0% |
| 6 | Borno | Bama | Rural | 50 | 18.0% |
| 7 | Yobe | Bade | Urban/Semi-urban | 50 | 54.0% |
| 8 | Yobe | Yusufari | Rural | 50 | 24.0% |
| 9 | Adamawa | Jada | Rural | 50 | 26.0% |
| 10 | Adamawa | Fufore | Semi-urban | 50 | 28.0% |
| 11 | Taraba | Takum | Semi-urban | 50 | 46.0% |
| 12 | Plateau | Jos East | Semi-urban | 50 | 52.0% |
| 13 | Plateau | Jos South | Urban | 50 | 66.0% |
| 14 | Benue | Gboko | Urban/Semi-urban | 50 | 58.0% |
| 15 | Nasarawa | Awe | Rural | 50 | 26.0% |
| 16 | Nasarawa | Keffi | Urban | 50 | 62.0% |
| 17 | Niger | Bida | Urban | 50 | 64.0% |
| 18 | Niger | Mokwa | Semi-urban | 50 | 52.0% |
| 19 | Kwara | Ifelodun | Semi-urban | 50 | 46.0% |
| 20 | Kwara | Ilorin West | Urban | 50 | 70.0% |
| 21 | Kogi | Lokoja | Urban | 50 | 68.0% |
| 22 | Kogi | Okene | Urban/Semi-urban | 50 | 60.0% |
| 23 | FCT | Bwari | Urban/Semi-urban | 50 | 64.0% |
| 24 | Kaduna | Kagarko | Rural | 50 | 26.0% |
| 25 | Kaduna | Lere | Semi-urban | 50 | 28.0% |
| 26 | Katsina | Kankia | Rural | 50 | 22.0% |
| 27 | Katsina | Malumfashi | Semi-urban | 50 | 48.0% |
| 28 | Zamfara | Tsafe | Rural | 50 | 24.0% |
| 29 | Zamfara | Zurmi | Rural | 50 | 20.0% |
| 30 | Sokoto | Gwadabawa | Rural | 50 | 24.0% |
| 31 | Sokoto | Yabo | Semi-urban | 50 | 28.0% |
| 32 | Kebbi | Jega | Semi-urban | 50 | 54.0% |
| 33 | Kebbi | Yauri | Semi-urban | 50 | 48.0% |
| 34 | Kano | Kabo | Rural | 50 | 26.0% |
| 35 | Kano | Nassarawa | Urban | 50 | 70.0% |
| 36 | Jigawa | Malam Madori | Rural | 50 | 22.0% |
| 37 | Jigawa | Birnin Kudu | Semi-urban | 50 | 44.0% |

**Table 3.2: Socio-Demographic Profile of Respondents (N = 1,850)**

| **Variable** | **Category** | **Frequency (n)** | **Percentage (%)** |
| --- | --- | --- | --- |
| Age Category | 18–24 years | 278 | 15.0% |
|  | 25–34 years | 702 | 38.0% |
|  | 35–44 years | 551 | 29.8% |
|  | 45–54 years | 238 | 12.9% |
|  | 55 years and above | 81 | 4.4% |
| Sex | Male | 1,018 | 55.0% |
|  | Female | 832 | 45.0% |
| Educational Level | No formal education | 370 | 20.0% |
|  | Primary education | 481 | 26.0% |
|  | Secondary education | 610 | 33.0% |
|  | Tertiary education | 389 | 21.0% |

**Table 3.3: Overall Knowledge and Attitude Distribution**

| **Variable** | **Category** | **Frequency (n)** | **Percentage (%)** |
| --- | --- | --- | --- |
| Knowledge Level | Good Knowledge (≥70%) | 747 | 40.4% |
|  | Poor/Insufficient Knowledge (<70%) | 1,103 | 59.6% |
| Attitude Category | Positive Attitude (≥70%) | 1,080 | 58.4% |
|  | Neutral Attitude (50–69%) | 360 | 19.5% |
|  | Negative Attitude (<50%) | 410 | 22.1% |

**Table 3.4: Association Between Settlement Setting and Polio Knowledge Level**

| **Setting** | **Good Knowledge** | **Poor Knowledge** | **Total** | **Chi-Square (χ²)** | **p-value** |
| --- | --- | --- | --- | --- | --- |
| Predominantly Rural | 178 (23.7%) | 572 (76.3%) | 750 (100.0%) | 144.00 | < 0.001 |
| Urban / Semi-urban | 569 (51.7%) | 531 (48.3%) | 1,100 (100.0%) |  |  |
| Total | 747 (40.4%) | 1,103 (59.6%) | 1,850 (100.0%) |  |  |

**Table 3.5: Primary Sources of Polio Immunization Information**

| **Information Source** | **Frequency (n)** | **Percentage (%)** |
| --- | --- | --- |
| Questionnaire | 995 | 53.6% |
| Health workers | 195 | 10.5% |
| Community & Traditional Leaders | 320 | 17.2% |
| Town Announcers | 220 | 11.9% |
| Religious Leaders | 120 | 6.5% |
| Total | 1,850 | 100.0% |

**4. DISCUSSION**

This multi-state assessment provides critical insights into the prevailing community knowledge and behavioral attitudes towards polio immunization across Northern Nigeria and the FCT. Although Nigeria was certified free of wild poliovirus in 2020, persistent cVDPV transmission underlines the fragility of population immunity when routine coverage lapses [2, 4].

A major empirical finding of this study is the substantial urban-rural disparity in polio knowledge. While 51.7% of respondents in urban and semi-urban LGAs possessed good knowledge, only 23.7% of rural respondents demonstrated adequate understanding (χ² = 144.00, p < 0.001). Rural communities in Northern Nigeria frequently face compound vulnerabilities: reduced access to formal health education, lower literacy rates, limited healthcare infrastructure, and geographic isolation [6, 11]. These factors foster an environment where vaccine misconceptions and safety fears can persist unchecked [13, 16].

Interestingly, despite significant knowledge deficits, overall community attitude towards polio vaccination was predominantly positive (58.4%). This suggests that willingness to accept vaccination often exists even where detailed scientific understanding of viral transmission is incomplete. However, a non-negligible minority exhibited neutral (19.5%) or negative (22.1%) attitudes. In-depth studies in northern communities have shown that non-compliance is strongly mediated by institutional distrust, perceived lack of tangible healthcare benefits, and religious concerns [8, 9, 13].

Health workers (53.6%) and radio broadcasts (10.5%) emerged as prominent channels of health communication alongside community leaders (17.2%). This highlights the indispensable role of trusted healthcare personnel and mass radio messaging in rural settings [16]. Traditional and religious leaders also accounted for a key portion of primary influence, confirming prior evidence that community leaders are vital gatekeepers for vaccine endorsement in Northern Nigeria [7, 10, 17].

**5. CONCLUSION AND RECOMMENDATIONS**

While community attitudes towards polio immunization are generally supportive across Northern Nigeria, significant gaps in disease knowledge persist, particularly in rural and hard-to-reach settlements. Addressing these gaps is vital to halting cVDPV transmission and sustaining high immunization coverage.

Based on the findings, the following policy recommendations are proposed:

- **Tailored Rural Risk Communication:** Develop context-specific health education packages delivered in local languages (Hausa/Fulani) targeting rural communities with low health literacy.
- **Leveraging Local Gatekeepers:** Systematically engage traditional rulers, religious leaders, and ward development committees to counteract vaccine misinformation and build community ownership.
- **Primary Healthcare Integration:** Bundle polio immunization campaigns with comprehensive maternal and child health packages (e.g., Vitamin A supplementation, deworming, routine childhood vaccines) to reduce campaign fatigue and increase perceived value.
- **Strengthening Microplanning:** Enhance GIS mapping and microplanning to reach zero-dose children in remote, riverine, nomadic, and security-compromised settlements.
- **Capacity Building for Frontline Workers:** Train community health extension workers (CHEWs) and vaccinators in interpersonal communication and empathetic counselling on vaccine safety.
- **Continuous Behavioral Surveillance:** Establish routine KAP tracking at the LGA level rather than relying solely on aggregated state-level indicators to catch emerging hesitancy early.

**REFERENCES**

1. World Health Organization. Poliomyelitis (Polio) Fact Sheet. Geneva: WHO; 2024.

2. World Health Organization Regional Office for Africa. Nigeria, WHO and partners reaffirm commitment to end all forms of polio by 2030. Brazzaville: WHO AFRO; 2025.

3. World Health Organization Regional Office for Africa. Nigeria launches large-scale vaccination campaign to protect 106 million children against measles, rubella and polio. Brazzaville: WHO AFRO; 2025.

4. World Health Organization. Statement of the Forty-fifth meeting of the Polio IHR Emergency Committee. Geneva: WHO; 2026.

5. National Primary Health Care Development Agency. Brief history and mandate. Abuja: NPHCDA; 2026.

6. National Population Commission (NPC) [Nigeria], ICF. Nigeria Demographic and Health Survey 2018. Abuja, Nigeria, and Rockville, Maryland, USA: NPC and ICF; 2019.

7. Nasir SG, Aliyu GG, Ya’u I, Kalbarczyk A, Saleh MA, Imam A, et al. A model for improving the quality of oral polio vaccine campaigns in Kano State, Nigeria. BMC Public Health. 2016;16:1180.

8. Yahya M. Polio vaccines—difficult to swallow: the story of a controversial campaign in Nigeria. IDS Working Paper 261. Brighton: Institute of Development Studies; 2007.

21. Global Polio Eradication Initiative. Polio programming in Nigeria is addressing gender-related barriers to reach more children with immunization. Geneva: GPEI; 2026.

**APPENDIX I: COMMUNITY QUESTIONNAIRE**

**Title: Community Knowledge and Attitude Towards Polio Immunization in Northern Nigeria
Instruction:** This questionnaire is for academic research purposes. Do NOT write your name. Participation is entirely voluntary and confidentiality is guaranteed.

**SECTION A: SOCIO-DEMOGRAPHIC INFORMATION**

1. Age: [ ] 18–24 years	[ ] 25–34 years	[ ] 35–44 years	[ ] 45–54 years	[ ] 55+ years

2. Sex: [ ] Male	[ ] Female	[ ] Prefer not to say

3. Marital Status: [ ] Single	[ ] Married	[ ] Divorced/Separated	[ ] Widowed

4. Highest Educational Level: [ ] None	[ ] Qur’anic only	[ ] Primary	[ ] Secondary	[ ] Tertiary

5. Occupation: [ ] Farmer	[ ] Trader	[ ] Civil Servant	[ ] Health Worker	[ ] Artisan	[ ] Student	[ ] Other

6. State: ____________________	7. LGA: ____________________	8. Community: ____________________

9. Residence Classification: [ ] Rural	[ ] Semi-urban	[ ] Urban

**SECTION B: KNOWLEDGE OF POLIO AND IMMUNIZATION**

10. Have you heard of poliomyelitis (polio)? [ ] Yes	[ ] No

11. Is polio a preventable disease? [ ] Yes	[ ] No	[ ] Don't Know

12. Can polio infection cause permanent paralysis in children? [ ] Yes	[ ] No	[ ] Don't Know

13. Can poliovirus spread from person to person? [ ] Yes	[ ] No	[ ] Don't Know

14. Is vaccination an effective way to protect children from polio? [ ] Yes	[ ] No	[ ] Don't Know

15. Have you heard of the Oral Polio Vaccine (OPV drops)? [ ] Yes	[ ] No

16. Does a child need more than one dose of polio vaccine to be fully protected? [ ] Yes	[ ] No	[ ] Don't Know

17. Is polio vaccination still important when there are no active cases in the community? [ ] Yes	[ ] No	[ ] Don't Know

18. Can a child receive additional polio drops during campaigns even if previously vaccinated? [ ] Yes	[ ] No	[ ] Don't Know

19. Can good sanitation and hygiene help reduce the spread of polio? [ ] Yes	[ ] No	[ ] Don't Know

20. What is your PRIMARY source of information on polio vaccination?
 [ ] Health Worker	[ ] Radio	[ ] Television	[ ] Community/Religious Leader	[ ] Family/Friends	[ ] Town Announcer	[ ] Social Media

**SECTION C: ATTITUDE TOWARDS POLIO IMMUNIZATION**

Please rate your level of agreement with the following statements (SA = Strongly Agree, A = Agree, U = Undecided, D = Disagree, SD = Strongly Disagree):

| **Statement** | **SA** | **A** | **U** | **D** | **SD** |
| --- | --- | --- | --- | --- | --- |
| 21. Polio vaccination effectively protects children against serious paralysis. |  |  |  |  |  |
| 22. I trust health workers who administer polio vaccines in our community. |  |  |  |  |  |
| 23. Polio vaccines provided by public health authorities are safe and beneficial. |  |  |  |  |  |
| 24. I actively encourage other parents and caregivers to vaccinate their children. |  |  |  |  |  |
| 25. Community and traditional leaders should advocate for polio vaccination. |  |  |  |  |  |
| 26. Religious leaders should support and encourage child immunization. |  |  |  |  |  |
| 27. Repeated door-to-door vaccination campaigns are necessary for child health. |  |  |  |  |  |
| 28. I am willing to allow my eligible child to receive polio drops anytime vaccinators visit. |  |  |  |  |  |
| 29. I am concerned that polio vaccines might cause harm or infertility. |  |  |  |  |  |
| 30. I would refuse vaccination if a community member warned me against it. |  |  |  |  |  |
| 31. Vaccination campaigns should be combined with general primary healthcare services. |  |  |  |  |  |
| 32. I feel comfortable asking health workers questions about vaccine safety. |  |  |  |  |  |

**APPENDIX II: INFORMED CONSENT TEMPLATE**

**Study Title:** Community Knowledge and Attitude Towards Polio Immunization in Northern Nigeria
**Principal Investigator:** Muhammad Auwal Gidado (Department of Public Health, College of Health Technology Ningi)

**Purpose & Voluntary Participation:** You are invited to participate in a research study assessing community knowledge and attitudes regarding polio immunization. Participation is entirely voluntary. You may choose not to answer any question or withdraw at any point without penalty.

**Confidentiality:** No personal identification numbers, names, or addresses will be recorded on the questionnaire. All data will be aggregated and used strictly for academic and public health research purposes.

**Consent Statement:** I have read/been read the details of this study and voluntarily consent to participate.

Participant Signature / Thumbprint: ____________________	Date: ______________
Interviewer Signature: ____________________	Date: ______________

**APPENDIX III: VARIABLE CODING SCHEME**

| **Variable Name** | **Data Type** | **Coding Values** |
| --- | --- | --- |
| sex | Categorical | 1 = Male, 2 = Female |
| education | Ordinal | 0 = None, 1 = Qur'anic only, 2 = Primary, 3 = Secondary, 4 = Tertiary |
| residence | Categorical | 1 = Rural, 2 = Semi-urban, 3 = Urban |
| knowledge_item | Binary | 1 = Correct response, 0 = Incorrect / Don't know |
| attitude_item | Ordinal | 1 = Strongly Disagree to 5 = Strongly Agree (Reverse items inverted) |
| overall_knowledge | Binary | 1 = Good Knowledge (≥70%), 0 = Poor Knowledge (<70%) |
| overall_attitude | Binary | 1 = Positive Attitude (≥70%), 0 = Neutral/Negative (<70%) |
| accept_vaccine | Binary | 1 = Yes / Willing, 0 = No / Hesitant |
